# Determinants and outcomes in children with cochlear implants in Chile: A longitudinal study in the Latin American context

**DOI:** 10.1101/2025.11.19.25340553

**Authors:** Mario Bustos-Rubilar, Merle Mahon, Fiona Kyle

**Author notes:** Corresponding author. Division of Psychology and Language Sciences, Brain Faculty, University College London, 2 Wakefield St, London WC1N 1PJ.

## Abstract

Deafness from birth presents a major challenge to communication development and carries significant public health implications. Cochlear implants (CI) are a key intervention for those with severe to profound deafness. In Chile and across Latin America, policies exist to support early CI access. However, delays in implementation and limited monitoring reduce their effectiveness. As a result, variability persists in spoken language outcomes for deaf children using CI. This study followed 49 Chilean children who received a CI between 2017 and 2019, using hospital records and two parental surveys over a 12-month period. Language outcomes and progress were investigated using validated tools (CAP-II, SIR, CDI Vocabulary), alongside five selected key factors: additional disabilities, age at implantation, daily CI use, socioeconomic factors, parental training and confidence. Findings indicated a modest yet uneven improvement in speech perception and language abilities over the study period. Fewer than one-third of children used only spoken language at home. Auditory performance and speech intelligibility improved slightly, while receptive vocabulary scores plateaued, and vocabulary production increased modestly after one year of CI use. Daily CI use and parental training were the strongest predictors of language outcomes . Despite current/recent policy advances, CI outcomes continue to be limited by delayed access, fragmented services, and socioeconomic inequality. These results underline the need for tailored strategies to strengthen early intervention and language support in Chile and across Latin America.

## 1. Introduction

The global incidence of severe to profound deafness or hearing loss (HL) from birth affects over 430 million people worldwide (WHO, 2021). In Latin America, there are approximately 16 million children under the age of 15 who are deaf (OMS, 2018). In Chile, deafness affects 2.8 per 1,000 newborns, with prevalence among school-aged children ranging from 0.2% to 7.8% (Torrente, Tamblay, Herrada, & Maass, 2023). Deafness usually refers to varying degrees of hearing loss. Deaf children, especially those born with severe to profound HL, often face major challenges in acquiring spoken language without early intervention. Without proper access to sign language and other communication supports, their cognitive development, social inclusion, and educational outcomes are significantly impacted. The economic cost of unaddressed childhood deafness in the region is high, with estimates ranging from USD 750 to 790 billion (WHO, 2017).

Cochlear implant (CI) is a electronic device used to support spoken language development in deaf children with severe and profound levels of deafness. Among the various factors influencing CI success, timely and structured early intervention is critical. This includes newborn screening by 1 month, diagnosis by 3 months, and hearing aid (HA) fitting by 6 months (Yoshinaga-Itano, Baca, & Sedey, 2010). Early CI implantation and switch-on, ideally between 9 and 12 months, has been shown to result in better spoken language outcomes (Park, Gagnon, & Brown, 2021). However, despite the existence of such protocols in several Latin American countries, delays remain common due to systemic and socioeconomic barriers, which diminish the effectiveness of interventions.

At the policy level, CI programmes are in place in countries such as Mexico, Argentina, and Chile (CSG México, 2010; Ministerio de Salud, 2016; MINSAL, 2013). Nonetheless, monitoring is challenging and coordination between sectors is often lacking. In Chile, there have been notable advances: the inclusion of hearing loss diagnosis and treatment in the public system in 2013 (MINSAL, 2016); CI coverage through a high cost policy law in 2018; and the launch of a national universal newborn hearing screening programme in 2020 (Minsal, 2022). Despite these developments, significant implementation gaps persist, including delayed diagnosis, geographical inequities, socioeconomic burdens on families, and a non-integrated framing of deafness between social and medical approaches.

A recent national observational study (Bustos-Rubilar, Kyle, & Mahon, 2025) found that this unfavorable context, including socio-economic disparities, age at CI implantation, borough development, variability in CI use, and parental engagement, may influence speech perception as part of spoken language outcomes. They reported that later implantation, and socio-economic factors contributed to wide variability and lower speech perception performance; for example, only 12% achieved good levels of speech comprehension and 38% were rated as having unintelligible connected speech. This is in stark contrast to more advanced speech perception outcomes typically reported in international studies from high-income countries with children implanted early (e.g. Niparko et al. 2010).

Spoken language development has been the primary outcome targeted by CI in high-income countries for the past three decades (Ganek, McConkey Robbins, & Niparko, 2012). Early research focused on speech perception, with consistent evidence showing that implanted children significantly improved phoneme recognition and intelligibility. For instance, a systematic review found that within one to three years, children with CI could identify tones and recognise words in noise (Y. Chen, Wong, Zhu, & Xi, 2017). More recently, attention has shifted to broader language competencies such as vocabulary, comprehension, and pragmatics (Ching & Dillon, 2013; Crowe & Dammeyer, 2021; Duchesne & Marschark, 2019). Language outcomes for CI children outcomes vary widely, influenced by factors such as aetiology, additional disabilities, age at switch-on, socioeconomic status, service access, parental education, and daily CI use (Yoshinaga-Itano, Sedey, Wiggin, & Mason, 2018).In the UK, only 36% of children with CI achieved age-appropriate language parameters compared to 81% of hearing peers (RCSLT & NDCS, 2017). In Australia, the Longitudinal Outcomes of Children with Hearing Impairment (LOCHI) study of 451 children reported a mean global standardised language score of 77.8 versus the normative 100 at age three (Ching, Dillon, Leigh, & Cupples, 2018). Similarly, a US longitudinal study showed significant yearly gains in language reception and expression, though average scores remained below age expectations even after several years (Niparko et al., 2010).

Despite reported improvements, many deaf children with CI continue to lag behind their hearing peers even in high income contexts (Ching et al., 2018). Most studies rely on small, English-speaking samples, often excluding children with multiple disabilities (Crowe & Dammeyer, 2021), and data from low- and middle-income countries,where over 80% of children with hearing loss live, remain limited (Queiroz, Bevilacqua, & Costa, 2010; Rayes, Al-Malky, & Vickers, 2019). Longitudinal studies in high-income countries have provided valuable insights into key factors influencing better spoken language outcomes including early age at intervention, high family socioeconomic status, good quality of parent-child interaction, and good maternal education level (Ching et al., 2018; Niparko et al., 2010; Persici et al., 2022; Yoshinaga-Itano et al., 2018).

There is a pressing need for evidence that reflects more diverse social and health system realities such as those found in low- and middle-income countries. Similar research to that already conducted in high income countries is urgently needed in Chile and Latin America to monitor and evaluate spoken language development among children with CI. The Bustos-Rubilar et al. (2025) study reported significant delays in speech perception among deaf children with CI in Chile, but the impact of this upon their language outcomes and trajectories is unknown. Thus, the current study follows up 49 of the 107 children who participated in the Bustos-Rubilar et al. (2025) study with the aim of tracking their spoken language progress over twelve months and identifing key factors that predict spoken language outcomes in Chilean children with cochlear implants, contributing to the broader Latin American context.

## 2. Methods

We conducted a longitudinal study that followed the initial national characterisation study by Bustos-Rubilar, Kyle, and Mahon (2025), which provided an audit of hospital clincal record records for an initial data baseline of potential participants. Thereafter, parents whose children with CI met the inclusion criteria were invited to take part in the current study. We conducted an online parental survey, between September 2020 to March 2021 at two time points, 12 months apart. Thus, we gathered data from two main sources: 1) Hospital clinical records from children with CI attending public hospitals in Chile (Appendix 1), and 2) the online survey (Apprendix 2) completed by each of the parent/closest caregiver of each child.

Two Research Ethics Committees approved this study: Faculty of Medicine, University of Chile (167-2020) and University College London (UCL) (LCD-2020-13). The study was registered with UCL Data Protection, and all approved data management and transfer procedures were followed. The methods were pre-registered on the OSF platform (https://osf.io/jeyg6). Written informed parental consent was completed for each respondent at the beginning of the online survey.

A bilingual committee of English and Spanish speakers developed protocols and parent surveys, adapted them for online use and distribution via email or text. To ensure accessibility, we included sign language interpreters, video calls, and telephone facilitators as needed (Bustos-Rubilar, Kyle, Tapia-Mora, Hormazábal-Reed, & Mahon, 2022)

### Participants

From participants in the aforementioned observational study (Bustos-Rubilar, Kyle, & Mahon, 2025) , we invited all parents and caregivers of children aged two to six years who had received their first CI at a Chilean public hospital between January 2017 and December 2019 and had used the device for at least one year prior to joining the study. Children also needed to have complete clinical data, with parents or caregivers able to participate in follow-up evaluations. Of the 153 children initially identified by the Chilean Ministry of Health, 107 took part in the observational study (Bustos-Rubilar, Kyle, & Mahon, 2025), 62 met the initial inclusion criteria for the current study , and 49 gave consent to take part. The final sample of 49 deaf children with CI represents 79% of children implanted in Chile at that time.

At Time 1, children ranged in age from 2 years 7 months to 6 years 9 month at T1 with a mean age of 5 years 3 months (SD=13.5 months). The average age of CI implantation for participating children was 2 years and 8 months (SD = 10.2 months), ranging from 12 months to 4 years and 6 months; the median age of CI implantation was 2 years and 10 months.

Socioeconomic status data, based on Socioeconomic Health Insurance (SHI) classification, revealed that 34.7% of the children were from low-income households and 14.3% from low-middle income. For caregiver education (FHE), defined as the closest caregiver to the child but not necessarily the most educated in the family, 44.9% had completed secondary school and 20.4% held a university degree. Most children (75%) had no additional developmental difficulties and rich spoken language environment at school.

### Materials

Parent surveys and clinical records:We collected data on background demographics, audiological and rehabilitation factors for participating children through a combination of hospital clinical records and from parent surveys completed at Time 1 and Time 2 (see data in Tables 1 and 2).

**Table 1.**
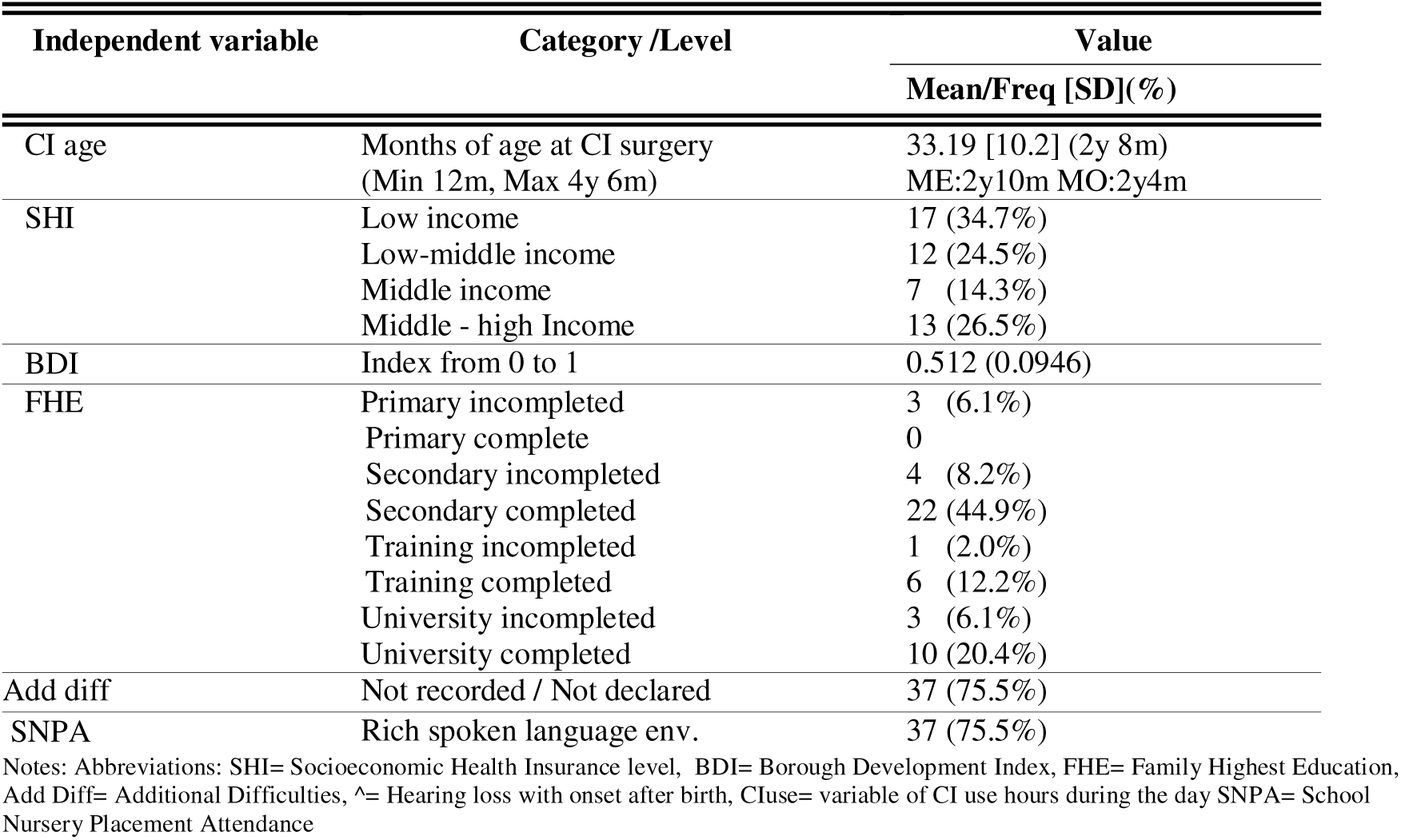
Background demographics and audiological variables collected at Time 1 only.

| Independent variable | Category /Level | Value |
| --- | --- | --- |
|  |  | Mean/Freq [SD](%) |
| CI age | Months of age at CI surgery<br>(Min 12m, Max 4y 6m) | 33.19 [10.2] (2y 8m)<br>ME:2y10m MO:2y4m |
| SHI | Low income | 17 (34.7%) |
|  | Low-middle income | 12 (24.5%) |
|  | Middle income | 7 (14.3%) |
|  | Middle - high Income | 13 (26.5%) |
| BDI | Index from 0 to 1 | 0.512 (0.0946) |
| FHE | Primary incompleted | 3 (6.1%) |
|  | Primary complete | 0 |
|  | Secondary incompleted | 4 (8.2%) |
|  | Secondary completed | 22 (44.9%) |
|  | Training incompleted | 1 (2.0%) |
|  | Training completed | 6 (12.2%) |
|  | University incompleted | 3 (6.1%) |
|  | University completed | 10 (20.4%) |
| Add diff | Not recorded / Not declared | 37 (75.5%) |
| SNPA | Rich spoken language env. | 37 (75.5%) |

Notes: Abbreviations: SHI= Socioeconomic Health Insurance level, BDI= Borough Development Index, FHE= Family Highest Education, Add Diff= Additional Difficulties, ^= Hearing loss with onset after birth, CIuse= variable of CI use hours during the day SNPA= School Nursery Placement Attendance

**Table 2.** Descriptive statistics for sociodemographic, audiological and rehabilitation variables for children with CI at Times 1 and 2.

| Independent variable | Category /Level | Time 1 | Time 2 |
| --- | --- | --- | --- |
|  |  | Mean/Freq [SD](%) | Mean/Freq[SD](%) |
| Age | Chronological age years/months<br>(T1:Min 2y7m, Max 6y9m)<br>(T2:Min 3y7m, Max 7y11m) | 5 y 3 m [SD=13.5m]<br>ME: 5y7m MO:3y 9m | 6 y 3 m [SD=12.5]<br>ME: 6y8m MO:6y 9m |
| Gender | Male | 27 (55.1%) | NR |
| CI status | Unilateral CI w/o contra HA | 35 (71.4%) | 29 (59.2%) |
|  | Unilateral CI with contra HA | 10 (20.4%) | 10 (20.4%) |
|  | Bilateral CI | 4 (8.2%) | 10 (20.4%) |
| CI condition | Functioning-Without issues | 37 (75.5%) | 35 (71.4%) |
|  | Func. but with some issues | 9 (18.4%) | 11 (22.4%) |
|  | Not Func. – Tech. issues | 1 (2.0%) | 1 (2.0%) |
|  | Not in use | 2 (4.1%) | 2 (4.1%) |
| Rehabilitation attendance | Yes, Face to Face | 33 (67.3%) | NR |
|  | Yes, Hybrid | 8 (16.3%) | NR |
|  | Yes, Online | 6 (12.2%) | NR |
|  | No | 2 (4.1%) | NR |
| Easy rehabilitation attendance | Yes | 36 (73.5%) | NR |
|  | No | 13 (26.5%) | NR |
| Commute time | Very short time | 1 (2.0%) | NR |
|  | Short Time | 11 (22.4%) | NR |
|  | Enough Time | 13 (26.5%) | NR |
|  | Long Time | 8 (16.3%) | NR |
|  | Very long Time | 7 (14.3%) | NR |
|  | Not attending | 9 (18.4%) | NR |
| Education attendance | Not attending any ed. | 6 (12.2%) | 5 (10.2%) |
|  | Special school for DC | 6 (12.2%) | 7 (14.3%) |
|  | Mainstream ed. w/o SNA | 2 (4.1%) | 3 (6.1%) |
|  | Mainstream ed. with SNA | 29 (59.2%) | 24 (49.0%) |
|  | Special school for SLD | 5 (10.2%) | 0 (0%) |
|  | Mainstream nursery | 1 (2.0%) | 10 (20.4%) |
| CI use (ord) | Never | 2 (4.1%) | 2 (4.1%) |
|  | Sometimes | 1 (2.0%) | 3 (6.1%) |
|  | Frequently | 5 (10.2%) | 4 (8.2%) |
|  | Always | 41 (83.7%) | 40 (81.6%) |
| CI hrs per day | From min=0.0 max=19.0 hrs.<br>*CIuse= $\mu$ T1,T2= 10.6 [3.3] | 10.2 [3.6] | 11.2 [3.7] |
| Behavioural problems | No | 36 (73.5%) | NR |
|  | Yes | 13 (26.5%) | NR |
| Frequency rehabilitation | Weekly | 37 (75.5%) | 33 (67.3%) |
|  | Each 2 weeks | 7 (14.3%) | 8 (16.3%) |
|  | Monthly | 3 (6.1%) | 6 (12.2%) |
|  | Less than once per month | 2 (4.1%) | 2 (4.1%) |
| Parental engagement | No | 2 (4.1%) | 14 (28.6%) |
| • CI Training | Yes | 47 (95.9%) | 35 (71.4%) |
| Parental engagement | No confidence | 1 (2.0%) | 2 (4.1%) |
| • CI Confidence | Very Poor Confident | 1 (2.0%) | 2 (4.1%) |
|  | Poor Confident | 3 (6.1%) | 5 (10.2%) |
|  | Somewhat Confident | 22 (44.9%) | 18 (36.7%) |
|  | Very Confident | 22 (44.9%) | 22 (44.9%) |

Notes: Abbreviations: Freq= Frequency, [SD]= Standard deviation, NR= Not recorded, T1= Time 1, T2= Time 2, , CI= Cochlear Implant, Tech.= Technical, \*, HA= Hearing aid/s, SNA= Special Needs Assistance, SLD= Speech and Language Disorders, CI Training: Parental Engagement variable about previous parental training with the CI, CI Confidence: Parental Engagement variable about confidence with the CI intervention.

Speech perception and spoken language outcomes:

Parents completed five assessments of their child’s speech perception and language ability as part of the parent surveys at Time 1 and at Time 2:

1. Speech perception ability was assessed using the Chilean Version of the Categories of Auditory Performance CAP II (CAPII). The Chilean CAPII is a nine-point rating scale that had been previously adapted into a parent reported assement of speech perception (see Bustos-Rubilar et al., 2022).
2. Speech production was measured using the Chilean Version of the Speech Intelligibility Rating Scale SIR (SIR). The SIR is a five-point rating scale previously adapted into a parent reported scale (see Bustos-Rubilar et al., 2022).
3. Receptive vocabulary ability (CDI RV) was assessed by parents completing the understanding checklist of the Chilean Communicative Development Inventory: Words and Gestures (CDI) (Farkas & Klein, 2010)
4. Expressive vocabulary ability (CDI EV) was assessed by parents completing the production checklist of Chilean Communicative Development Inventory: Words and Gestures (CDI) (Farkas & Klein, 2010)
5. Parents were asked to indicate the communication used at home.

See Table 3 for further information about the CAPII, SIR and communication used at home variable.

**Table 3.** Descriptive statistics for speech perception and spoken language outcomes in children with CI at Times 1 and 2.

| Type of communication at home |  |  |  |  |
| --- | --- | --- | --- | --- |
|  | T1 |  | T2 |  |
|  | Freq. | % | Freq. | % |
| Mixed (Sign and Spoken Language) | 34 | 69.4 | 24 | 49 |
| Spoken Language | 12 | 24.5 | 14 | 28.6 |
| Other (babbling, early communication, gestures) | 2 | 4.1 | 9 | 18.4 |
| Sign Language | 1 | 2 | 2 | 4.1 |
| Chilean Spanish CAP-II Categories |  |  |  |  |
|  | T1 |  | T2 |  |
|  | Freq. | % | Freq. | % |
| 0. No awareness of environmental sounds | 3 | 6.1 | 5 | 10.2 |
| 1. Awareness of environmental sounds | 0 | 0 | 2 | 4.1 |
| 2. Response to speech sounds | 3 | 6.1 | 1 | 2 |
| 3. Recognition of environmental sounds | 4 | 8.2 | 2 | 4.1 |
| 4. Discrimination of at least two speech sounds | 4 | 8.2 | 8 | 16.3 |
| 5. Understanding of common phrases without lipreading | 12 | 24.5 | 13 | 26.5 |
| 6. Understanding of conversation without lipreading with a familiar talker | 2 | 4.1 | 4 | 8.2 |
| 7. Use of a telephone with a familiar talker | 11 | 22.4 | 2 | 4.1 |
| 8. Understanding/Following group conversations. | 6 | 12.2 | 7 | 14.3 |
| 9. Use the telephone with an unknown speaker in an unpredictable context. | 4 | 8.2 | 5 | 10.2 |

Chilean Spanish SIR Categories
|  | T1 |  | T2 |  |
| --- | --- | --- | --- | --- |
|  | Freq. | % | Freq. | % |
| 1. Connected speech is unintelligible. Pre-recognisable words in spoken language. | 23 | 46.9 | 16 | 32.7 |
| 2. Connected speech is unintelligible; intelligible speech is developing in single words. | 3 | 6.1 | 8 | 16.3 |
| 3. Connected speech is intelligible to a listener who concentrates and lip-reads. | 3 | 6.1 | 2 | 4.1 |
| 4. Connected speech is intelligible to a listener who has little experience of a deaf person's speech. | 15 | 30.6 | 15 | 30.6 |
| 5. Connected speech is intelligible to all listeners. The child is understood easily in everyday contexts. | 5 | 10.2 | 8 | 16.3 |

CDI scores
|  | T1 |  | T2 |  |
| --- | --- | --- | --- | --- |
|  | Mean | SD | Mean | SD |
| CDI Receptive Vocabulary (RV) (Min= 0, Max= 160) | 102.77 | [56.04] | 103.10 | [63.49] |
| CDI Expressive Vocabulary (EV) (Min= 0, Max= 160) | 37.14 | [48.41] | 53.53 | [56.41] |

Notes: In Communication at Home "Mixed Language" includes spoken and sign language, "Other" includes any other predominant communication at home. Abbreviations: GeersM= Geer and Moog Latin American Categories of Speech Perception, CAP II= Categories of Auditory Performance, SIR= Speech Intelligibility Rating Scale Categories.

### Data analysis strategy

Descriptive statistics (frequencies, medians, means, quantiles, SDs) were conducted for all demographic and audiological variables and spoken language outcome assessments.

Inferential analyses were used to examined the associations between the two language outcomes assessed in our study: CDI Receptive Vocabulary (RV) and CDI Expressive Vocabulary (EV) at both timepoints, and five key factors known from previous research to influence language development in children with CI:

(1) The presence of additional difficulties; (2) Age of implantation (CIAge); (3) CI use; (4) Parental engagement including CI Training and CI Confidence; and (5) Social economic determinants of health which included Socioeconomic Health Insurance (SHI and the Borough Development Index (BDI) – this is a Chilean index used as a proxy for social determinants of health showing well-being and access to services in boroughs (Hernández Bonivento et al., 2020).

A multivariate analysis was used to evaluate which of these factors predicted progress in spoken language over 12 months. In addition, the child’s age, Family Education, and School Nursery Placement Attendance (SNPA) were included Data visualisations were created to show the association between the individual progression made in language outcomes from time 1 to time 2 and the presence of additional difficulties, CI Age and CI use.

Inferential statistics included Pearson/Spearman correlations, t-tests, ANOVA, ANCOVA, and other relevant parametric or non-parametric tests, with significance set at p < .05. To control for Type I errors while preserving statistical power, Bonferroni corrections set a stricter alpha of 0.0028. Findings with p-values between .05 and .0028 were flagged as potentially significant. We examined collinearity among our included variables; variables with a variance inflator factor (VIF)>5 were removed. All statistical analyses were performed using R Studio version 1.4.

## 3. Results

### 3.1 Descriptive stats for independent variables and evaluated outcomes

We categorised participants based on their geographical residence location and sociodemographic characteristics which were plotted on a map of Chile, with each national borough shaded according to a Chilean BDI. Figure 1 illustrates the geographical distribution of our sample by Chilean boroughs using the BDI scores (from 0.0 to 1.0). Most of the children came from the central area (53%), specifically Santiago (31%), and southern regions (33%). Santiago and Concepcion have the highest BDI scores (0.78 and 0.64, respectively), whereas the remaining areas ranged below 0.60. The average BDI value of our sample was 0.51 compared to the average BDI of the country as a whole which is 0.37 . This suggests that children with CI in this study came from slightly more developed boroughs than the average in the country.

**Figure 1.**
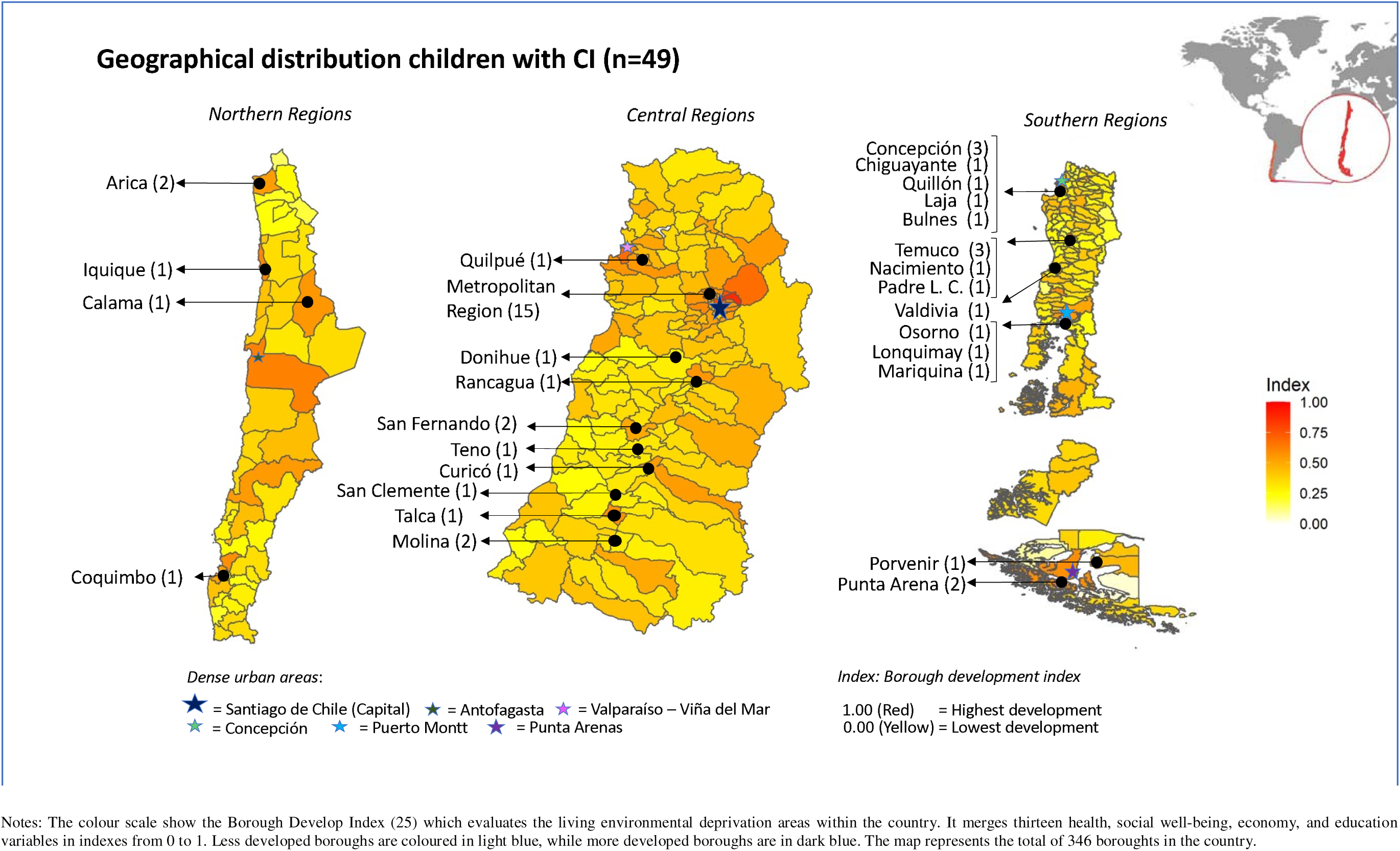
Geographical distribution of deaf children 228 with CI (N=49) by BDI in Chile.

Table 2 contains descriptive statistics for all sociodemographic, audiological and rehabilitation variables for the sample at both T1 and T2. Most participants had a unilateral cochlear implant (CI) without a contralateral hearing aid (HA) at both T1 (71.4%) and T2 (59.2%). The majority had a functioning CI at both T1 (75.5%) and T2 (71.4%) and attended face-to-face rehabilitation at T1 (67%). Most caregivers (73.5%) reported easy access to rehabilitation.

In terms of education, most children were enrolled in mainstream schools with special needs support at T1 (59.2%) and T2 (49.0%), with spoken language being the dominant mode of communication (75.5%). CI usage was consistently high, with over 80% of children always using their devices and the average daily usage was 10.6 hours. Behavioural issues related to CI use were minimal, with 73.5% reporting no problems. Weekly rehabilitation attendance was common at T1 (75.5%) and T2 (67.3%). Regarding parental engagement, the majority had received CI training at T1 (95.9%) and T2 (71.4%), and most caregivers reported being somewhat or very confident in managing the device at both time points.

The descriptive statistics for the five speech perception and spoken language outcome measures for children at both T1 and T2 are shown in Table 3. Mixed communication (spoken and sign language) was the most commonly reported mode of communication at both time points—69% at T1 and 49% at T2—followed by spoken language only (25% at T1, 29% at T2). A Chi-squared test showed no significant change over time, X²(3, N = 49) = 6.67, p = .083, with a small effect size (Cohen’s d = -0.31, 95% CI [-0.71, 0.09]). Auditory performance (CAP-II) scores had a median score at 5^th^ category at both T1 and T2 (range: 0–9) and there was no significant change over time, Wilcoxon T = 333.5, p = 0.093. The fifth category, *‘Understanding of common phrases without lipreading”,* was the most frequently reported (12 children at T1, 13 children at T2). While 17 children improved, 5 showed a decline of more than two categories. No). For speech intelligibility (SIR), the median rose from 2 to 3 showing no statistically significant improvement from T1 to T2 (Wilcoxon T=79.5, p=0.212), and Category 1, reflecting unintelligible connected speech, was still the most frequently reported (47% at T1, 33% at T2).

Vocabulary results from CDI RV and CDI EV showed stable understanding (Time 1 mean RV = 102.77 (SD=56.04) and Time 2 mean RV =103.10 (SD=63.49) and an increase in vocabulary production (Time 1 mean EV=37.14 (SD=48.41) and Time 2 mean EV=53.53 (SD = 56.41)). In the CDI RV results, a ceiling effect was observed in 16 children (32%). ANOVA results revealed a significant subtest main effect, F(1,193) = 49.323, p < .0001, but no significant effect of time (F = 0.853, p = .357) or interaction (F = 0.780, p = .378).

Despite overall stable or positive trends across most of the outcome variables between T1 and T2, it is important to note that 14 children showed a decline on at least one of the speech perception and language outcomes, including CAPII, CDI-RV, and CDI-EV, which may seem contradictory considering typical child development A review of the characteristics of this subgroup revealed that most of these children used mixed or alternative communication, and five had additional developmental conditions, including two with autistic spectrum disorder. Additionally, challenges in literacy for the self-completion of the survey could have affected the responses from some parents, especially considering that only one parent in this subgroup of children had completed post-secondary education. These potentially anomalous results could have influenced the overall results of the sample. However, we conducted the analyses both with and without this subgroup, and similar main results were found and therefore we decided to include all participants.

### 3.2 Statistical analysis between key factors and language outcomes

A series of analyses were conducted to examine specific associations between five key factors known from previous research to influence language development in children with CI, and the two language outcomes assessed in our study: CDI Receptive Vocabulary (RV) and CDI Expressive Vocabulary (EV) at both timepoints

1. The presence of additional difficulties Two two-way ANOVAs (with Bonferroni-adjusted alpha threshold) revealed that children without additional difficulties had significantly better language outcomes than those with additional difficulties at both T1 and T2. For the CDI RV, there was a significant main effect of additional difficulties (F(1, 95) = 16.642, p <.0001), but no main effect of Time (F(1, 95) = 0.012, p = .912), or interaction between additional difficulties and time (F(1,95)= 0.0064, p=.800). Similarly for CDI EV, there was a main effect of additional difficulties (F(1, 95) = 13.778, p <.0001), and no main effect of Time (F(1, 95) = 0.389, p = .534). The two-way interaction between additional difficulties and time was also not significant (F(1,95)= 0.000, p=.984). Two further ANCOVA tests were run to determine if children without additional difficulties make more progress on the CDI RV and CDI EV than children with additional difficulties. After controlling for T1 score results, the interaction between additional difficulties and outcome results at T2 showed no significant difference in CDI RV results (F (1,46) = 0.000407, p= 0.984) and CDI EV results (F(1,46) = 0.591, p= .446).
2. Age of CI implantation Two two-way ANOVAs were run to evaluate if children implanted before 24 months of age had better-spoken language outcomes on CDI RV and CDI EV than children implanted after 24 months. For the CDI RV, there was no significant main effect of age of CI implantation (F (1, 95) = 0.607, p = .438) and no main effect of time of evaluation (F (1, 95) = 0.010, p = .919), and no significant two-way interaction between the age of CI implantation and time of evaluation (F(1,95)= 0.545, p=.502). Similarly, for CDI EV, there was no main effect of age of CI surgery (F(1, 95) = 0.300, p = .585), no main effect of time of evaluation (F(1, 95) = 0.340, p = .561) and no significant two-way interaction between additional difficulties and time (F(1,95)= 0.608, p=.437). Finally, two ANCOVA tests were run to determine if children implanted before 24 months made more progress in spoken language between T1 and T2. Before Bonferroni-adjusted alpha thresholds were applied, children implanted before 24 months showed significantly greater gains in CDI EV over time (F (1,46) = 1.746 p = .049), but not in CDI RV (F(1,46)= 4.521, p = .52).
3. CI daily use in more or less than 8 hours/day Two two-way ANOVAs investigated whether children who used their CI for more than 8 hours per day had better scores in both receptive and expressive vocabulary as measured by the CDI RV and CDI EV at both timepoints. There was a significant effect of CI use in CDI RV (F(1,95) = 29.052, p < .001) and CDI EV (F(1,95) = 23.003, p < .001). However, there were no significant main effect of time in CDI RV ( F(1,95) = 0.014, p = .907), CDI EV (F(1,95) = 0.419, p = .519), and no significant interaction between CI use and time in CDI RV (F(1,95) = 0.052, p = .821) and CDI EV (F(1,95) = 0.001, p = .973). An ANCOVA was also conducted to examine the effect of CI hours of use on vocabulary gains over time, controlling for baseline (T1) scores. After adjustment for T1 results, the analysis showed no significant difference in CDI RV scores (F(1,46) = 0.033, p = .856). However, there was a significant effect of CI use on CDI EV gains at T2 (F(1,46) = 5.49, p = .004), but this effect did not remain significant after applying the Bonferroni correction for multiple comparisons
4. Social economic determinants of health Proxy variables related to social determinants were analysed in association with spoken language outcomes. Using two-way ANOVAs, BDI as a binary variable (<0.5 N=19, >0.5, N=30) showed a significant association with CDI RV (F(1,95) = 6.65, p = .03) but this was no longer significant after applying the Bonferroni correction. There was no significant in and CDI EV (F(1,95) = 4.01, p = .07). In a correlation test, there was a low but positive correlation between BDI and CDI EV at T1 (r = .37, p = .009) suggested that participants from higher BDI areas scored slightly better, though this association weakened after statistical adjustment. In contrast, socioeconomic health insurance level (SHI) and family highest education (FHE) showed no significant main effects or interactions with time on either CDI RV (SHI= F(1,46) = 0.141, p = .709, FHE= (F(1,46) = 1.404, p = .242)) and EV (SHI= F(1,46) = 0.018, p = .849, FHE= (F(1,46) = 0399, p = .531)). Similarly, after using ANCOVA to assess prediction of vocabulary progression from Time 1, none of these social variables predicted progression between timepoints after controlling for baseline performance.
5. Parental engagement At T2, CI confidence showed a moderate positive correlation at with CDI RV (rho = 0.40, p = .005) and a potential correlation with CDI EV (rho = 0.322, p = .023). There was no significant effect of CI training on CDI RV (F(1,95) = 1.520, p = .22) and CDI VP (F(1,95) = 0.960, p = .33), nor did it influence progress over time using ANCOVA when adjusting for baseline scores in CDI RV (F(1,46) = 0.011, p = .918) and CDI VP (F(1,46) = 2.912, p = .950).

We created paired boxplots to visually depict the associations between the presence of additional disabilities, CI age, and hours of CI use and CDI RV and CDI EV at both timepoints, and the individual growth in vocabulary between the two time points (see Figure 2).

**Figure 2.**
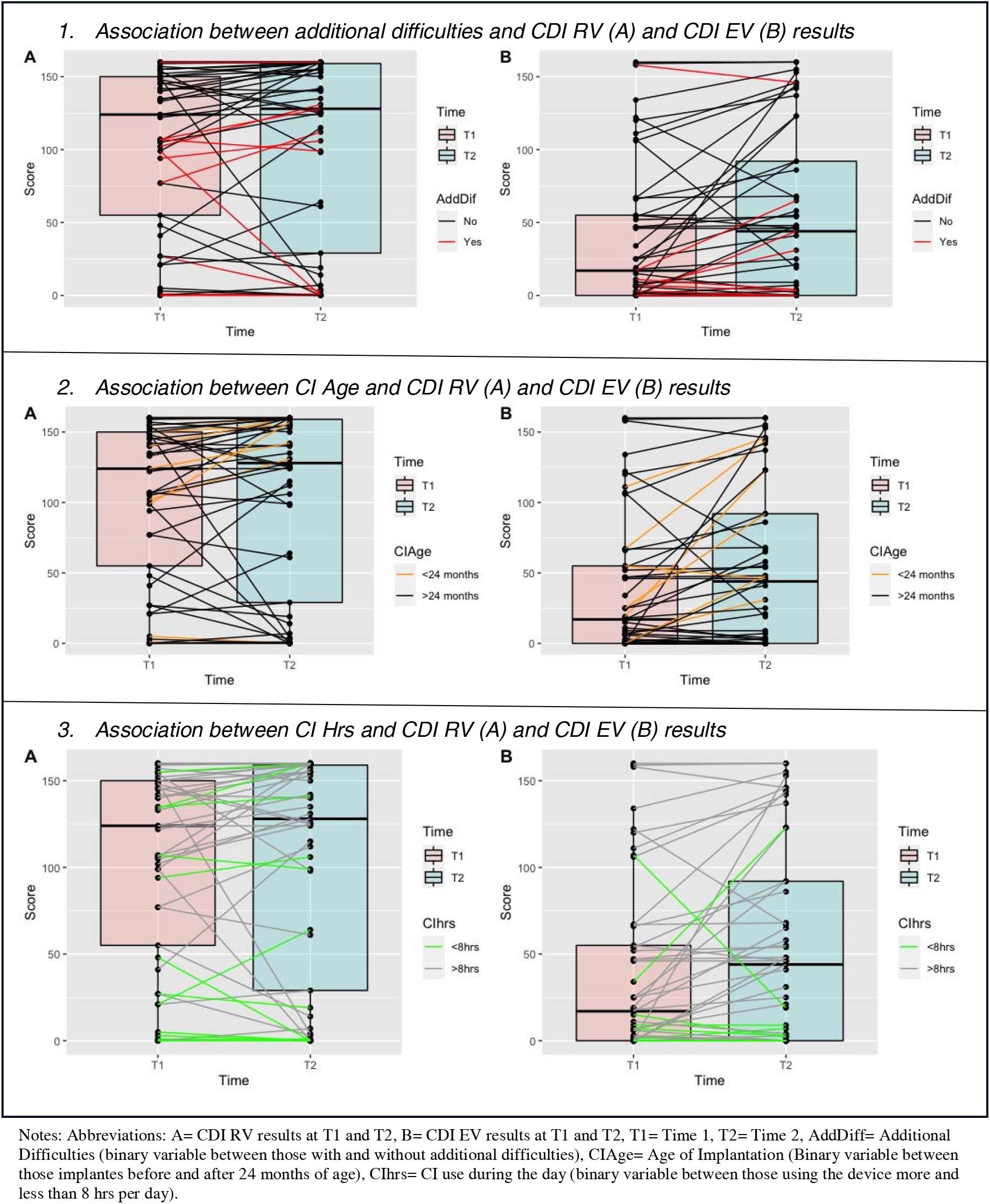
Individual progression between Time 1 and time 2 in (A) CDI RV and (B) CDI EV considering three factors.

A multivariate linear regression model is presented in Table 4. Independent variables included in the model were those analyzed in the univariate analyses and identified as significantly associated with outcomes: presence of additional difficulties, parental CI confidence, daily CI use, and the richness of the spoken language environment in educational settings (SNPA). Table 4 displays two multivariate linear regression models for CDI RV and CDI EV. Higher CDI RV scores were positively associated with more hours of daily CI use (β = 6.77; 95% CI = 1.58–11.97; p = .012) and higher parental CI confidence, representing parental engagement (β = 19.95; 95% CI = 3.15–36.74; p = .021). The model explained 38% of the variance in CDI RV scores. Therefore, daily CI use and parental CI confidence were significant predictors of better CDI RV outcomes. In contrast, no significant predictors were identified for CDI EV scores.

**Table 4.** Multivariate regression analyses for CDI RV and CDI EV scores results and potential factors affecting spoken language outcomes at T2.

| *Predictors* | CDI RV results at T2 |  |  |  | CDI EV results at T2 |  |  |  |
| --- | --- | --- | --- | --- | --- | --- | --- | --- |
| | *Estimates* | $\beta$ | *CI*s | *p* | *Estimates* | $\beta$ | *CI* | *p* |
| (Intercept) | -80.78 | -185.91 | -166.84 – 5.29 | 0.065 | -48.51 | -111.64 | -131.58 – 34.57 | 0.246 |
| Add diff | -8.91 | -20.49 | -49.30 – 31.49 | 0.659 | -12.16 | -27.998 | -51.16 – 26.83 | 0.533 |
| CI Confidence | 20.79 | 47.85 | 6.03 – 35.55 | **0.007** | 7.86 | 18.08 | -6.39 – 22.11 | 0.272 |
| CI use | 6.77 | 15.58 | 1.58 – 11.97 | **0.012** | 3.97 | 9.14 | -1.04 – 8.99 | 0.117 |
| Less rich SL Ed | 36.69 | 84.45 | -4.65 – 78.04 | 0.081 | 39.84 | 91.70 | -0.07 – 79.76 | 0.050 |
| Observations | 49 |  |  |  | 49 |  |  |  |
| R<sup>2</sup> / R<sup>2</sup> adj | 0.381 / 0.325 |  |  |  | 0.270 / 0.203 |  |  |  |

Notes: Abb: CDI RV T1 = CDI Vocabulary understand at T1, CDI RV T2 = CDI Vocabulary understand at T2, Add Dif= Additional Difficulties (Binary variable yes/no), CI Confidence= Confidence with the CI in Likert scale as parental engagement variable. CI use = CI use in hrs using the average between T1 & T2 during the day, Less rich SL Ed= School Nursery Placement Attendance (SNPA) binary variable, which included rich and less rich spoken language communication environment in education. Categories. **Alpha**= p-value **0.05** ‘\*’

## 4. Discussion

This is the first longitudinal study of language outcomes in young deaf children with CI in Chile and within the Latin American region. The sample included 79% of deaf children implanted at in the public health system in Chile between 2017 to 2019. The findings showed limited improvement in auditory outcomes using CAP II and only half of children were rated as intelligible to experienced listeners in speech intelligibility ratings, after one year of using the CI. Almost 30% of children with CI used spoken language at home at T2, with no significant change over a 12 month period. This limited spoken language use highlights a gap between the current findings and previous evidence about better outcomes in spoken language communication after one year of CI use (Niparko et al., 2010).

For the main outcome of spoken vocabulary knowledge, results indicated that receptive vocabulary scores (CDI RV) remained stable between the two time points (T1: 102.77 and T2: 103.1), while expressive vocabulary scores showed a non-significant increase (T1: 37.14 and T2: 53.53), suggesting children made limited progress in expressive vocabulary after one year of using a cochlear implant.This contrasts with studies reporting rapid vocabulary growth in children with CI during the first year (Chilosi et al., 2013; Faes, Gillis, & Gillis, 2017; Fagan, 2015; Jung et al., 2020; Koşaner, Uruk, Kilinc, Ispir, & Amann, 2013). For instance, Fagan et al. (2015) found in 9 US children that CDI scores rose from a mean of 0.67 words at 4 months to 102.33 words at 12 months. Likewise, Koşaner et al. (2013) observed Turkish children gaining almost 100 words in understanding and production between 6–9 months post-implantation. The above findings underscore the need of being cautious when applying results from small sample studies or from non-representative, homogeneous populations, as these findings may not generalise well to broader or more diverse groups, particularly in lower or middle income countries.

Approximately 34% of participants achieved the maximum score for receptive vocabulary at both time points, limiting the sensitivity of this task to measure language development over time. In contrast, no ceiling effect was observed for expressive vocabulary ,suggesting it is a more reliable method for tracking developmental trajectories of language over longer time periods. The overall low mean scores in both domains suggest a concerning underperformance among children with CI in the Chilean context. Ramírez-Barba et al. (2022) conducted the only other study of CI language outcomes in Latin America, but it was based on only 14 children with CI in Mexico. They reported a expressive vocabulary mean of 207.3 words and a receptive vocabulary mean of 156.7 words. It is difficult to compare our results with their results due to an incongruency in their study where expressive vocabulary scores exceeded receptive vocabulary scores, a pattern that is unusual and not consistent with established language development principles, as receptive vocabulary typically develops earlier and remains larger than expressive vocabulary.

In terms of factors associated with spoken language outcomes, our data show that children with CI who have additional difficulties tend to make less progress in spoken language over time. This finding aligns with results from existing studies (e.g. Cejas, Hoffman, & Quittner, 2015; Persici et al., 2022). However, children with additional disabilities may still benefit from early CI, because CI have positive effects on other critical aspects such as behavioural regulation, school performance, and social functioning (Cejas, Hoffman, & Quittner, 2015). The current findings underscore the need for more tailored and inclusive research and interventions for CI children with additional difficulties. The present study focused only on spoken language development, leaving out the potential benefits of sign language, which other research shows can greatly assist children with complex needs (Knoors & Marschark, 2018).

Age at implantation is another evidence-based predictor of spoken language development; however we found nosignificant association in our sample. One possible reason is the delay in CI implantation in our study population. In the current study, the average age of CI implantation in Chile was 2 years and 8 months, higher than other high income countries worldwide (Bruijnzeel et al., 2017). Early implantation was associated with improved vocabulary production, consistent with previous studies (Dettman et al., 2016; McKinney, 2017). However, early CI implantation remains a challenge in countries with underdeveloped hearing screening infrastructure. In Chile, universal neonatal screening only began as a pilot in 2020 (MINSAL, 2022), far behind the “1-3-6” early intervention guidelines (Yoshinaga-Itano et al., 2018). Latin America, overall, shows vast disparities in screening coverage (Giugliani et al., 2022), highlighting systemic barriers to timely intervention. While diagnostic support from industry can play a role, long-term improvements require robust and coordinated public healthcare systems.

Although socioeconomic and contextual factors have previously been identified as associated with language development, in this study the Borough Development Index (BDI), Socioeconomic Health Insurance (SHI), and Family Highest Education (FHE) were not significantly associated . This may be due to limitations in sample diversity or variable design, particularly since FHE did not always reflect the primary caregiver’s role or influence. BDI was a significant factor in the previous observational study (Bustos-Rubilar, Kyle, & Mahon, 2025), and also appeared significant in this study before the Bonferroni correction. This potentially suggests that children from better-resourced areas receive more effective CI intervention, while those from low-BDI boroughs may face additional access barriers. The reasons may range from gaps in professional healthcare coverage to limited caregiver knowledge about deafness and early intervention. A WHO report (2021) confirms that both stigma and lack of provider training obstruct care pathways, especially in low- and middle-income countries.

Daily CI use over 8 hours emerged as a strong predictor of positive language outcomes in our study. Longer daily CI use was significantly associated with improved receptive vocabulary. In the final regression model, daily CI use and CI Confidence together explained 38% of the variability in CDI RV scores at T2; each additional hour of CI use predicted an increase of 7 understood words. These findings echo earlier work in Norway and the U.S. (Wie, Falkenberg, Tvete, & Tomblin, 2007; Wiseman, Warner-Czyz, Kwon, Fiorentino, & Sweeney, 2021). However, sustained CI use involves coordinated effort among families, educators, and healthcare teams (Chen, Castellanos, Yu, & Houston, 2020). Behavioural resistance and overreporting by caregivers (Wiseman et al., 2021) can obscure real usage patterns. Objective data logging, combined with family support, could improve adherence with using the device to ensure consistent daily use. Currently, Chilean policy lacks specific guidance on CI use duration (Ministerio de Salud, 2019), unlike the UK’s NHS, which promotes multidisciplinary monitoring (NHS England, 2023). Expanding such frameworks regionally in Latin America could enhance language outcomes for children with CI.

Parental engagement was assessed through two variables: CI Confidence and CI Training. Children with CI whose parents reported higher confidence with CI tended to achieve better outcomes in both receptive and expressive vocabulary. This aligns with prior findings highlighting the role of family involvement in intervention success (Noblitt, Alfonso, Adkins, & Bush, 2018). Additionally, Suskind et al. (2016) demonstrated that enhancing parental confidence through a home-visiting program led to significant improvements in adult-child language interactions. Conversely, CI training showed no significant association with outcomes, possibly due to limited structured support for caregivers in Chile. National guidelines lack dedicated parental training frameworks (MINSAL, 2013), unlike models such as Queensland’s parenting schools (Children’s Health Queensland, 2016). This highlights the need for integrated support systems that enable and encourage active parental involvement.

The findings of this research have significant implications for the design of interventions and policies for children with CI in Chile and would be applicable across Latin America. One of the study’s key observations is the high level of service coverage, with nearly all children in this longitudinal sample receiving some form of professional intervention. This is a notable strength when compared to countries in the region such as Ecuador, Guatemala, and Venezuela, where access to cochlear implants and rehabilitation services is more limited (Emmett et al., 2016). However, this apparent strength must be balanced with concerns about the quality and consistency of intervention, including the limited use of contralateral hearing aids shown by only 28% of the sample reported bilateral CI or unilateral CI with a contralateral aid at Time 1, which may negatively impact language outcomes. Also, this contrasts with the reality in high-income countries, where children usually use bilateral CIs (Ching, Dillon, Leigh, & Cupples, 2018).

From a policy perspective, the Chilean framework for CI intervention (Ministerio de Salud, 2018) provides a foundational structure, but lacks operational detail on critical aspects such as parental training, inclusive education, and strategies for children with additional needs. As argued by Dettman et al. (2016) and others, the creation of specific, evidence-based technical guidance is essential to formalise intervention pathways. Moreover, integrated policies that go beyond the health sector, linking education, family support, deaf community integration, access to sign language, and social protection systems such as disability benefits and inclusive tailored social services are required to reduce inequality and support sustainable outcome (Gama, 2016; World Health Organization, 2021)

As a potential implication, social protection emerges as a key enabler of equitable intervention. There is research that supports mechanisms such as direct economic support to caregivers, as implemented in Panama’s *Angel Guardian Programme*, which allows a parent to receive financial support during the intervention period (Ullmann, Atuesta, Rubio, & Cecchini, 2022). In Chile, linking CI intervention with broader initiatives like Chile Crece Contigo, a nationwide early childhood development support programme, and national health insurance programs could help to ensure more inclusive and equitable support (Bedregal, Torres, & Carvallo, 2014).

As a result of the world-wide Covid-19 pandemic in 2020, the research design changed from in-person to online data collection. While this ensured continuity, it introduced potential limitations related to the validity of self-reported survey data, as the majority of information came from parents and caregivers.This introduces potential bias, as supported by previous findings on the limitations of self-reported parental data in developmental research (Wickenden & Elphick, 2016). Clinical records, including standardised spoken language assessments (e.g., CAPII, SIR, and CDI), were used to partially triangulate the data, but further research should include inputs from educators and clinicians to improve the reliability and multidimensionality of outcome evaluations. Notably, the pandemic may have disrupted intervention continuity, particularly in the early phases of the longitudinal study. This context must be considered when interpreting findings on CI use, treatment attendance, and spoken language progress. The inclusion of a second data collection point post-pandemic helps to mitigate this limitation, yet longitudinal comparisons with pre-pandemic data from similar populations remain limited (Emmett et al., 2021). Finally, there were some differences in the predictors of receptive and productive vocabulary, and these need to be explored further in future studies with larger sample sizes.

In conclusion, the current longitudinal study revealed distinct language outcome trajectories for children with CI in a Latin American context, with modest but uneven gains in varied outcomes including speech perception and language. The findings highlight persistent challenges to achieving stronger outcomes, including delayed implantation linked to late screening, fragmented rehabilitation services, limited parental training, and socioeconomic inequality. Importantly, daily CI use and parental confidence were the strongest predictors of receptive language progress, underscoring the central role of sustained family engagement alongside professional intervention. While Chile has established a robust policy framework and wide service coverage, concerns remain regarding quality and consistency. Strengthening early detection, embedding structured parental training, promoting inclusive education, and ensuring effective social protection measures are necessary to move from broad access to equitable outcomes.

## Acknowledgements

All authors attest they meet the ICMJE criteria for authorship and have reviewed and approved the final article. This article was supported by a full scholarship provided by the Chilean Government "Beca de Doctorado en el Extranjero Becas Chile, Convocatoria 2018, Ley N°21.053, Asociación Nacional de Investigación y Desarrollo (ANID)" and the Chilean Ministry of Health through the National Public Tender 757-89-l120.

## Ethics

Two Research Ethics Committees approved the study: The Faculty of Medicine, University of Chile (167-2020) and University College London (UCL) (RA040245/1). The approval considered data protection, procedures for collecting data and informed consent.

## COVID-19 acknowledgement

As a result of restrictions from March 2020, all measures were taken to follow the national guidance on research in health services

## Data availability and ethics

Data are available upon appropriate request.

